# A Web-Based Application for Real-Time Malaria Prediction using Environmental Variables

**DOI:** 10.1101/2025.08.20.25334055

**Authors:** Kumbirai Nicholas Matingo, Sydney Togarepi

## Abstract

Malaria remains a persistent public health challenge in Zimbabwe, particularly in rural districts such as Mudzi in Mashonaland East Province, where seasonal transmission and limited healthcare access undermine conventional control measures. This study presents the development and deployment of *MalariaDash*, a web-based application designed for real-time forecasting of malaria case counts using satellite-derived environmental data. The system integrates remote sensing inputs such as land surface temperature, vegetation indices, elevation, and proximity to water bodies with a Random Forest regression model trained on twenty-one months of health facility data. Predictor variables were selected based on their statistical significance in earlier modelling efforts.

Unlike existing studies that focus on retrospective risk mapping or static models, *MalariaDash* operationalises machine learning outputs into an interactive platform. The application dynamically retrieves environmental data through Sentinel Hub and weather APIs, enabling location-specific and time-specific malaria predictions. It is implemented using the Django web framework and provides a user interface where environmental conditions and predicted malaria incidence can be viewed for selected dates and areas.

This research demonstrates the practical value of linking environmental intelligence with predictive analytics in a rural African context. By delivering spatially explicit and near-real-time forecasts, *MalariaDash* enables health authorities to adopt proactive, targeted interventions rather than reactive responses. The approach illustrates a novel integration of geospatial modelling, machine learning, and operational web deployment aimed at improving local malaria control strategies.

**Author Summary:** In many rural parts of Zimbabwe, malaria continues to affect thousands of people every year. Although we know that changes in temperature, rainfall, and vegetation can influence where and when malaria outbreaks occur, local health workers often lack tools to make use of this information. In this study, we developed a web-based tool called MalariaDash that helps predict malaria cases using freely available environmental data from satellites. By combining these environmental signals with past malaria records, we created a system that allows users to select a location on a map and get a real-time forecast of malaria risk.

What makes this tool different is that it is not just a research model, but it is designed for practical use by local health teams. We tested it in Mudzi District, a rural area with a high malaria burden in Zimbabwe, and the system performed well in identifying high-risk times and places. Our hope is that this approach can help health workers respond earlier and plan better, especially in areas where resources are limited and time matters most.

## Introduction

Malaria remains one of Zimbabwe’s most pressing public health challenges, with significant incidence recorded annually, particularly in rural and low-lying districts such as Mudzi. The World Malaria Report (2024), notes that sub-Saharan Africa accounts for 94% of global malaria cases and 95% of related deaths, with Zimbabwe recording over 636,000 cases in 2023. This makes Zimbabwe one of the top 40 countries burdened by malaria (1). Despite long-standing interventions Ssuch as Indoor Residual spraying (IRS) and distribution of Insecticide-Treated Nets (ITNs), these strategies have shown diminishing returns in high-burden areas. This has mainly been attributed to operational delays, inconsistent coverage, and growing insecticide resistance (2,3).

The malaria situation in Mudzi District is particularly severe. Incidence rates are consistently higher than the national average, ranging from 111 to 264 cases per 1,000 people between 2016 and 2020. The district remains a persistent hotspot for transmission (4). Factors such as proximity to the Mozambique border, seasonal rainfall patterns, and socio-environmental conditions including stagnant water bodies, deforestation, and poverty exacerbate the risk. Limited access to healthcare, outdated paper-based reporting systems, and a lack of real-time spatial intelligence hinder the timely deployment of control measures (5,6).

While conventional malaria surveillance remains largely reactive, mobilising interventions only after case incidence rises, recent advances in remote sensing, geospatial analysis, and machine learning have enabled more proactive strategies. Environmental variables such as temperature, vegetation cover (NDVI), rainfall, elevation, and distance to water bodies are now well-established predictors of malaria transmission dynamics, particularly for Anopheles gambiae, the primary vector species in the region (7,8). However, translating these environmental insights into operational tools usable by local health authorities has remained a persistent gap in low-resource settings.

This study addresses that gap by developing and deploying MalariaDash, a web-based application for real-time malaria case prediction in Mudzi District. The platform integrates satellite-derived environmental data with a Random Forest regression model trained on facility-level case records. By allowing users to select locations and forecast periods, MalariaDash delivers location-specific risk predictions based on current and near-future environmental conditions.

This study contributes to the existing body of knowledge by identifying statistically significant environmental predictors of malaria within a hyper-local rural context. It develops a machine learning model trained on twenty-one months of epidemiological and environmental data, and subsequently operationalizes this model into a web-based application designed to support public health decision-making in Zimbabwe

Through this integrated framework, the research demonstrates how environmental intelligence can be leveraged for frontline disease surveillance and control planning.

## Materials and Methods

### Study Area

Mudzi District, located in the Mashonaland East province of Zimbabwe (Fig.1) near the Mozambique border, was selected as the study area due to its persistently high malaria incidence. Covering approximately 4,075 square kilometres of predominantly communal land, the district is characterised by low-lying valleys, seasonal water bodies, and highly variable rainfall patterns ranging from 600–800 mm annually (5).

**Figure 1:**
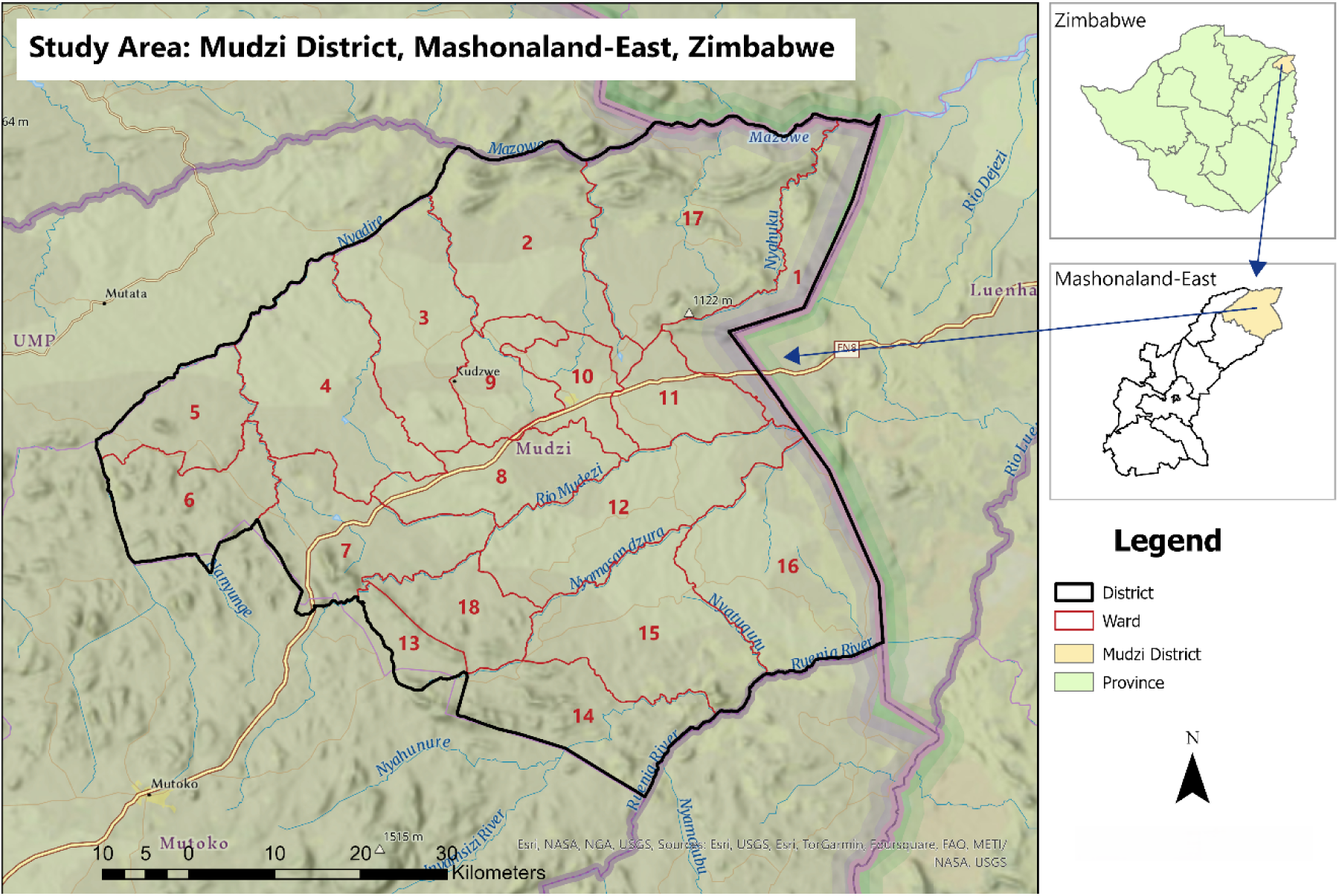
Study are map for Mudzi District.

The 2022 census recorded a population of 158,478, with most households’ dependent on subsistence farming and informal trade (9). The district is not included in Zimbabwe’s targeted malaria elimination zones, despite regularly ranking among the top five districts for malaria incidence between 2016 and 2020 (3).

### Data Sources

Malaria case data were collected from all 31 health facilities within the district for the period June 2023 to February 2025. Data were recorded monthly and accessed with ethical clearance from the Ministry of Health and Child Care (MoHCC). Variables were derived from remote sensing products and national datasets, as summarised in Table 1.

**Table 1:** Data and data sources.

| *Category* | *Variable* | *Data Source* |
| --- | --- | --- |
| Demographic | Number of households | GRID3, ZimStats Census data |
| Physical | Distance to:<br>- Water bodies and<br>- Health facilities | Computed via Euclidean distance |
| Environmental | Water bodies | Sentinel-2 Land use classification |
|  | Elevation (Altitude) | USGS |
|  | Vegetation (Normalized Difference Vegetation Index) | Sentinel-2 |
| Meteorological | Rainfall | The Climate Hazards and Centre (10) |
|  | Temperature (Land Surface Temperature) | MODIS |

Settlement data from GRID3 were used to delineate catchment zones for each health facility (Figure. 2), using a closest-facility network algorithm to account for patient mobility across administrative boundaries (11).

**Figure 2:**
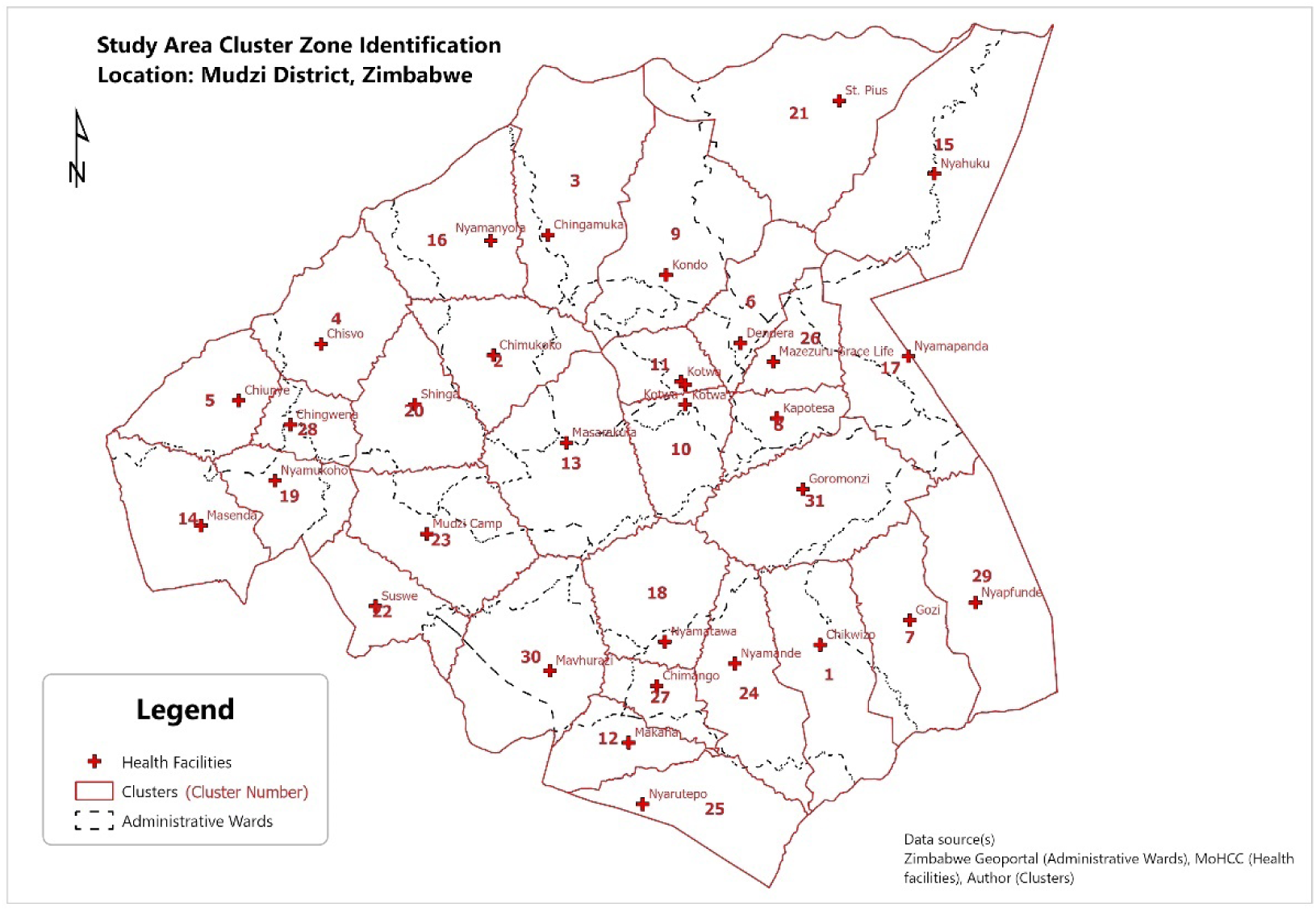
Cluster Zone identification map.

### Data Pre-processing

All environmental predictor variables were aggregated to monthly means and spatially averaged within each health facility’s catchment zone. Facility catchments were delineated using a network-based approach that considered travel distance to the nearest facility. This step ensured that predictor values accurately represented the environmental conditions influencing malaria transmission within each service area.

To capture delayed effects of environmental drivers on vector proliferation and parasite development, lagged variables were generated at one- and two-month intervals for key predictors, including temperature, NDVI, and precipitation. For example, a two-month lag for temperature reflects the combined incubation period of *Plasmodium falciparum* in mosquitoes and humans, as well as the larval development period. These lagged predictors were included alongside contemporaneous values in both statistical and machine learning models to test for time-delayed correlations.

Continuous predictors were normalised using a min-max scaling approach to standardise the variable ranges between 0 and 1, reducing biases from different unit scales and improving model interpretability. This pre-processing ensured that no single variable dominated the predictive model due to differences in magnitude.

### Model Development

A Random Forest regression model was constructed to predict monthly malaria case counts at the facility cluster level. Random Forest was chosen for its robustness to multicollinearity, ability to handle non-linear relationships, and superior performance in prior studies on disease prediction. The dataset was split into training (70%) and testing (30%) subsets, with stratification to preserve the temporal distribution of malaria cases.

Input variables included all environmental predictors outlined in Table 1, along with lagged versions of temperature, NDVI, and precipitation. Model hyperparameters such as the number of decision trees and maximum depth were optimised using a grid search approach with 10-fold cross-validation. Performance was evaluated using Root Mean Squared Error (RMSE) and the coefficient of determination (R²). Models such as linear regression, XGBoost, and Support Vector Regression were some of the models used for evaluation.

Variable importance analysis revealed that temperature (lag 2), precipitation (lag 1), and proximity to water bodies were among the strongest predictors of malaria incidence, providing interpretable evidence for the ecological relevance of these factors.

### Web Application Design

The trained Random Forest model was deployed in a web-based system named *MalariaDash*, designed to provide real-time malaria risk predictions. The application was built using the Django web framework (Python), leveraging its Model-View-Template (MVT) architecture to manage the interaction between the backend model, user interface, and dynamic data inputs.

The system is integrated with Sentinel Hub and weather APIs to automatically retrieve environmental data for user-selected locations and forecast dates. Users interact with a map-based interface to select a geographic point of interest and desired prediction period. Upon submission, the application dynamically processes environmental variables for the selected location, passes these inputs to the trained model, and outputs the predicted malaria case count along with associated environmental context.

**Figure 3.**
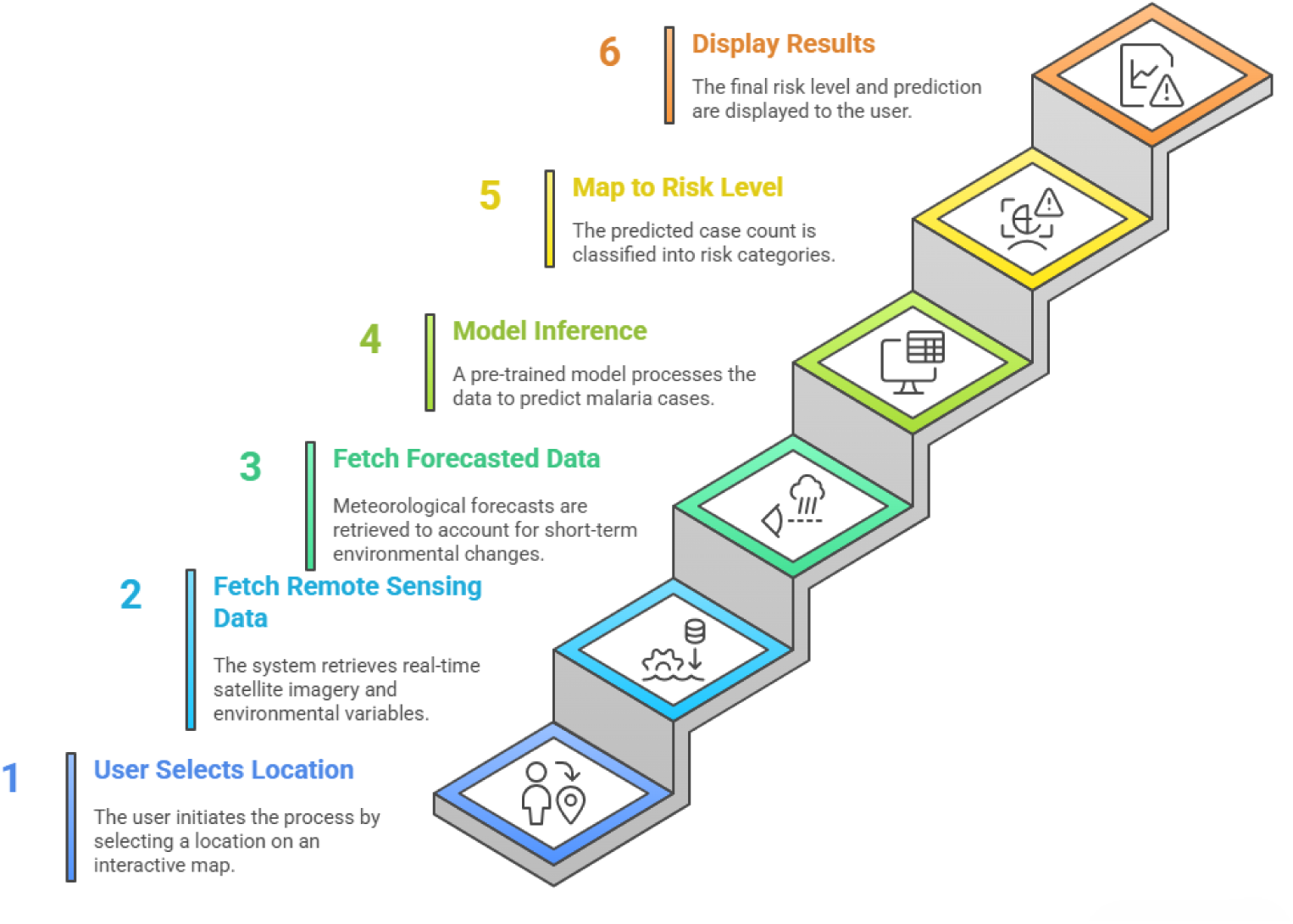
Workflow of data processing and prediction in MalariaDash.

A key feature of *MalariaDash* is its ability to store both predictions and associated environmental variables, creating a feedback loop for retraining and model improvement. The modular design of the application also allows future integration with national health information systems such as DHIS2, supporting both scalability and interoperability.

### Risk Mapping and Seasonal Analysis

To complement real-time forecasting, seasonal malaria risk maps were generated using ArcGIS Pro. Raster layers representing key predictors were normalised and combined using a weighted overlay model, where weights were derived from the statistical significance of variables in the regression analysis. Separate maps were produced for the wet (December– March), hot-dry (September–November), and cool-dry (April–August) seasons.

Risk scores were classified and visualised across both administrative wards and facility cluster zones, enabling multi-scale planning and intervention targeting.

## Results

### Environmental Determinants of Malaria Incidence

A twenty-one-month time series of malaria case counts across Mudzi District (Figure. 4) revealed seasonal fluctuations with notable peaks during and shortly after the rainy season. A sharp rise was observed between June and August 2023, tapering off through late 2024, and modestly increasing again in early 2025.

**Figure 4:**
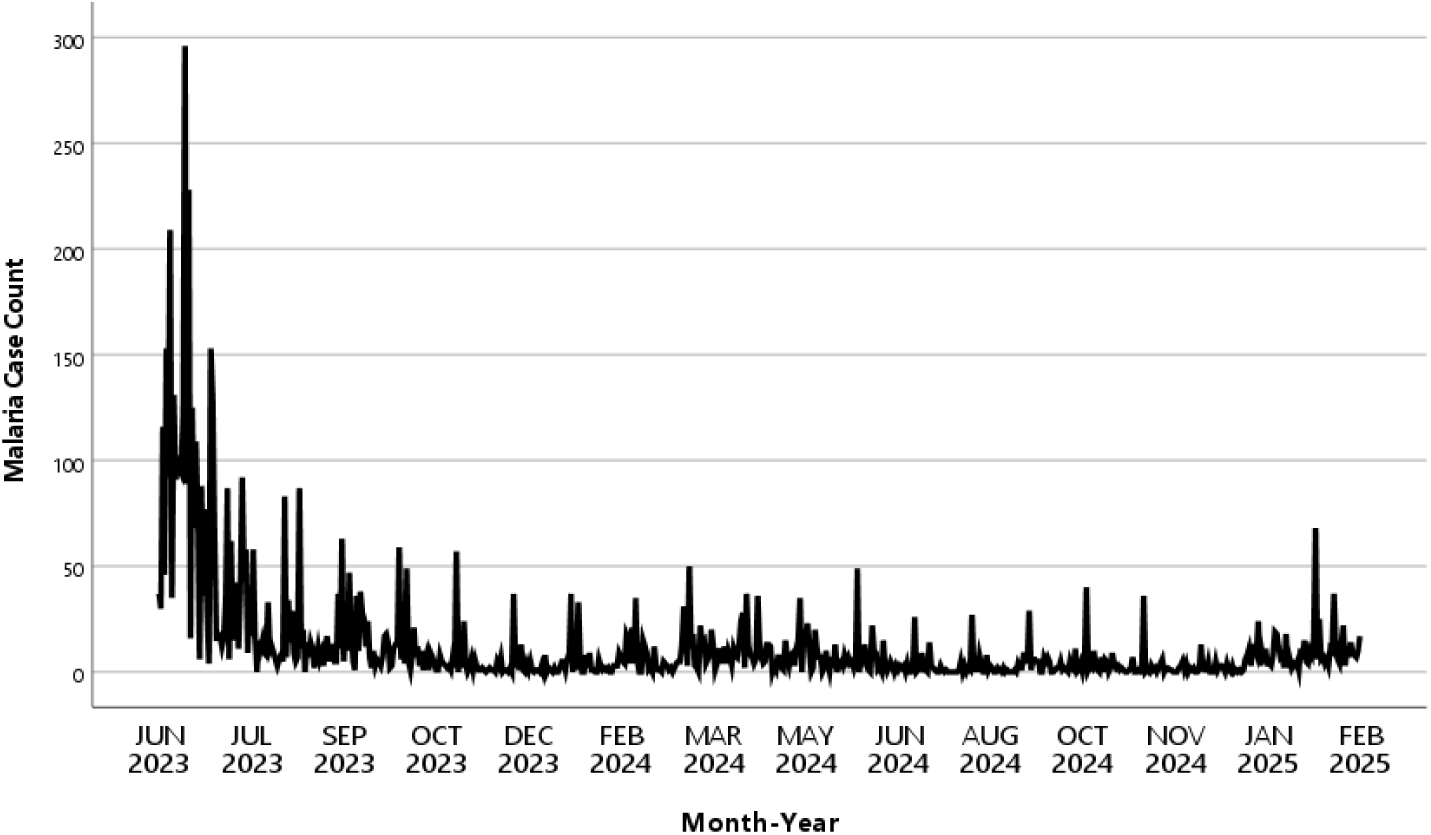
Time series malaria case counts across Mudzi district.

Initial modelling using Poisson regression exhibited overdispersion, with Pearson Chi-square and scaled deviance exceeding acceptable thresholds. A Negative Binomial Regression (NBR) model was therefore adopted. The NBR model identified several environmental predictors as statistically significant (p < 0.001), including land surface temperature, area of stagnant water bodies, distance to the nearest health facility, and distance to the nearest water body.

NDVI was not significant in isolation (p = 0.098), but it approached significance at Lag 2 and contributed meaningfully when included in interaction terms. Lagged analysis showed that both temperature and vegetation effects manifested with delays of one to two months, consistent with known vector development cycles.

Interaction effects were also tested. Notable findings included:

- Vegetation × Elevation exhibited a positive interaction (p = 0.003), indicating that vegetation effects vary by altitude.
- Precipitation × Elevation: Negative interaction (p < 0.001), suggesting that rainfall increases risk primarily in lower elevation areas.

Spatial autocorrelation testing via Global Moran’s I yielded non-significant results (Moran’s I = 0.0687, p = 0.449), implying that malaria incidence is not uniformly clustered. However, Local Moran’s I analysis identified specific outliers, including Low-High outlier clusters where lower incidence zones were surrounded by high-incidence areas possibly reflecting underreporting or targeted intervention success (Fig. 5).

**Figure 5:**
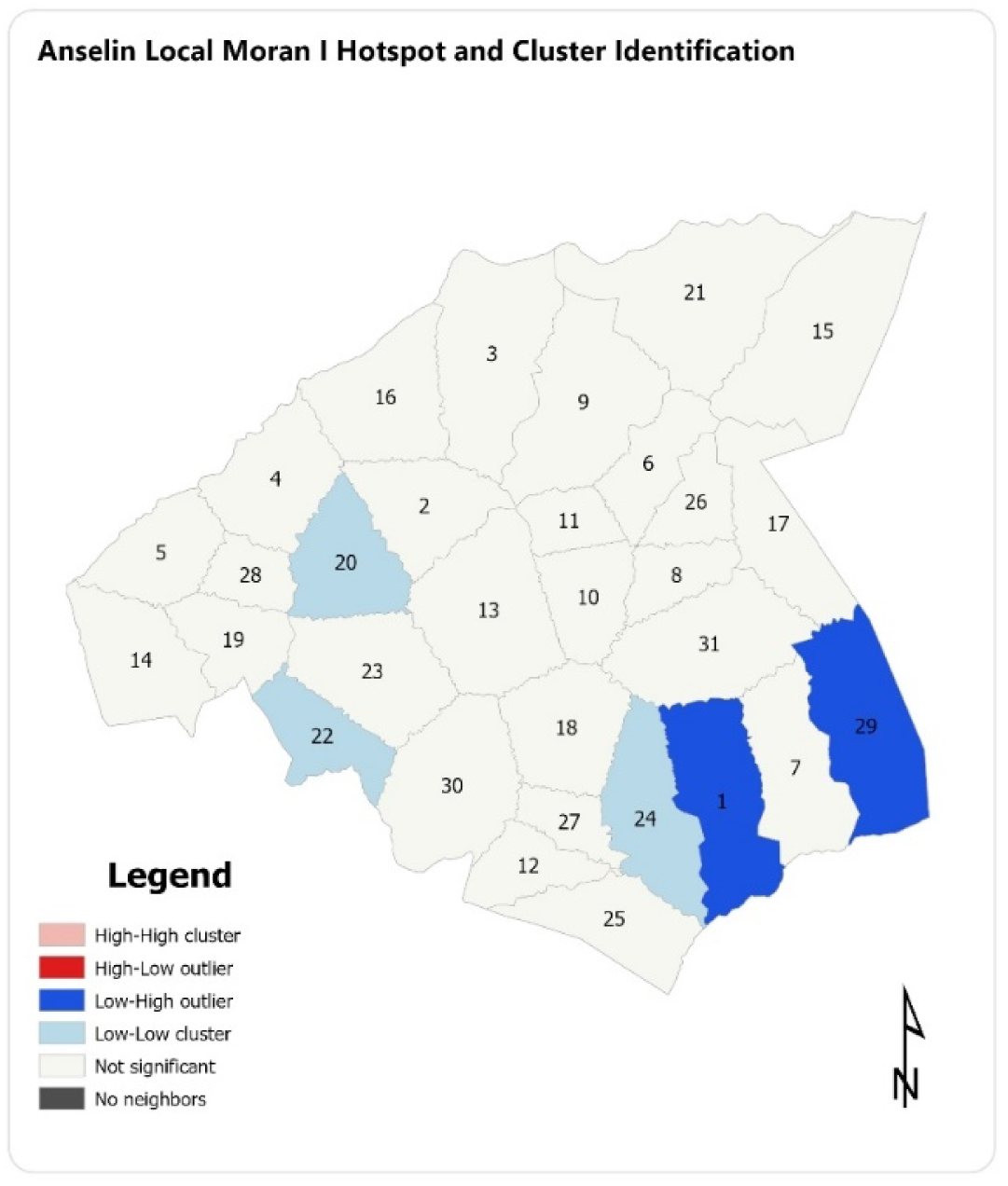
Anselin Local Moran ! Hotspot and Cluster Identification.

### Machine Learning Model Performance

A Random Forest regression model trained on environmental and spatial predictors outperformed all alternative models, including linear regression, XGBoost, and Support Vector Regression.

**Figure 6.**
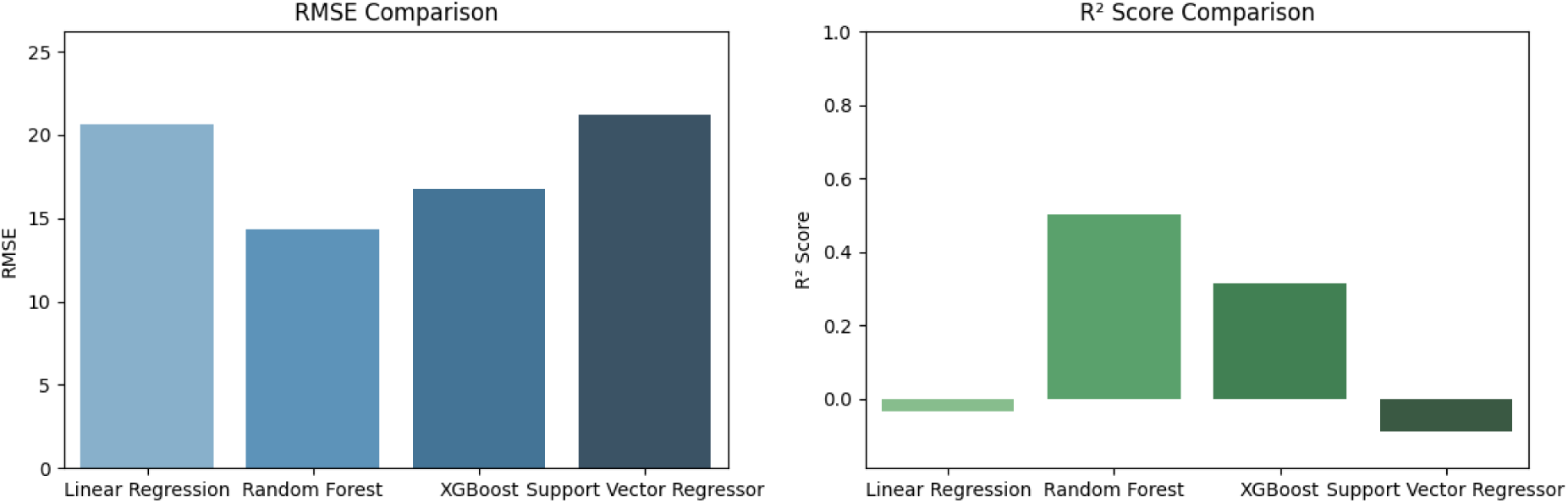

It achieved an RMSE of 14.33 and an R² of 0.502, indicating moderate predictive strength suitable for operational deployment.

**Figure 7.**
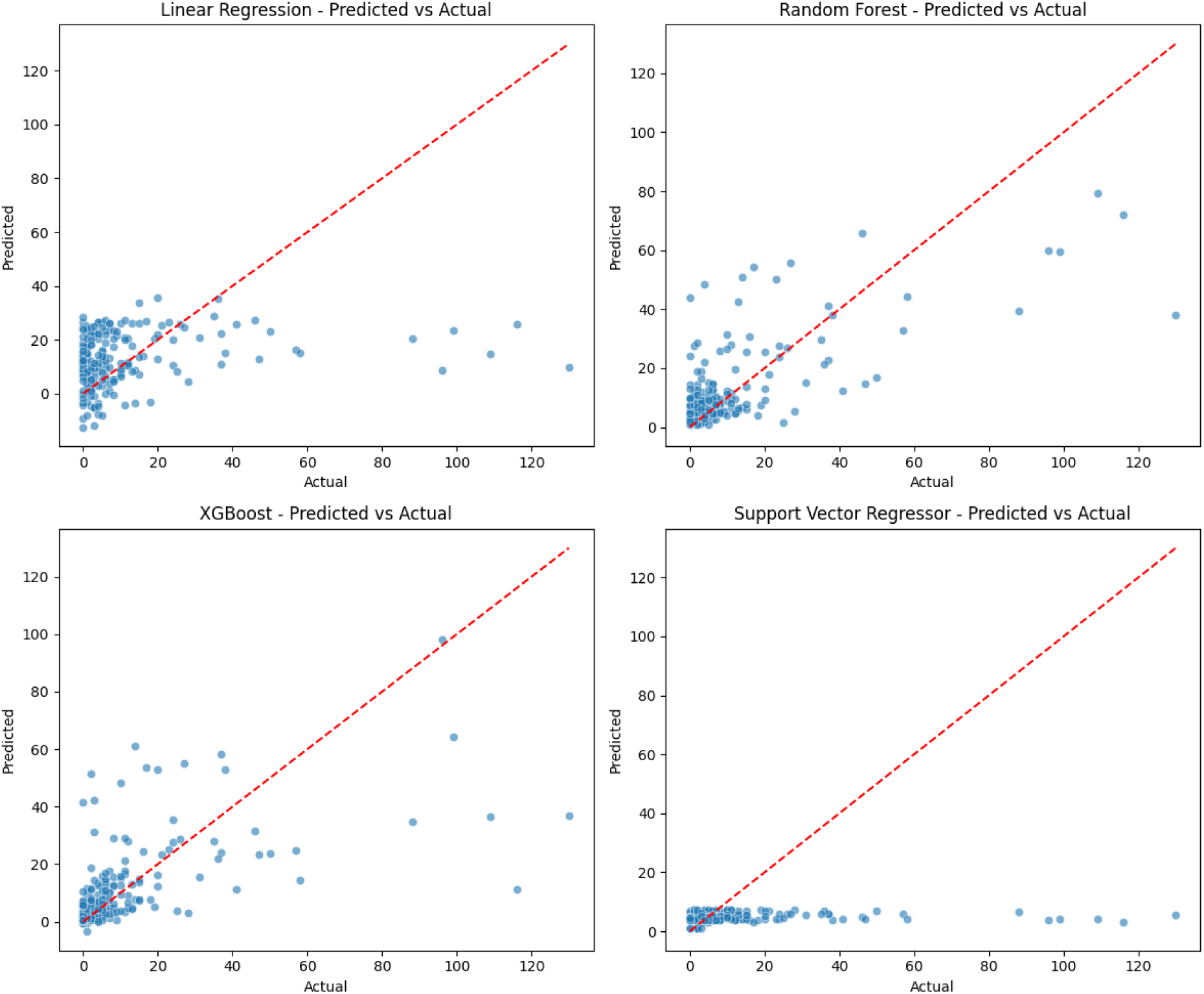

Residual analysis confirmed a balanced error distribution for the Random Forest model, while other models exhibited skewness and underestimation patterns.

**Figure 8.**
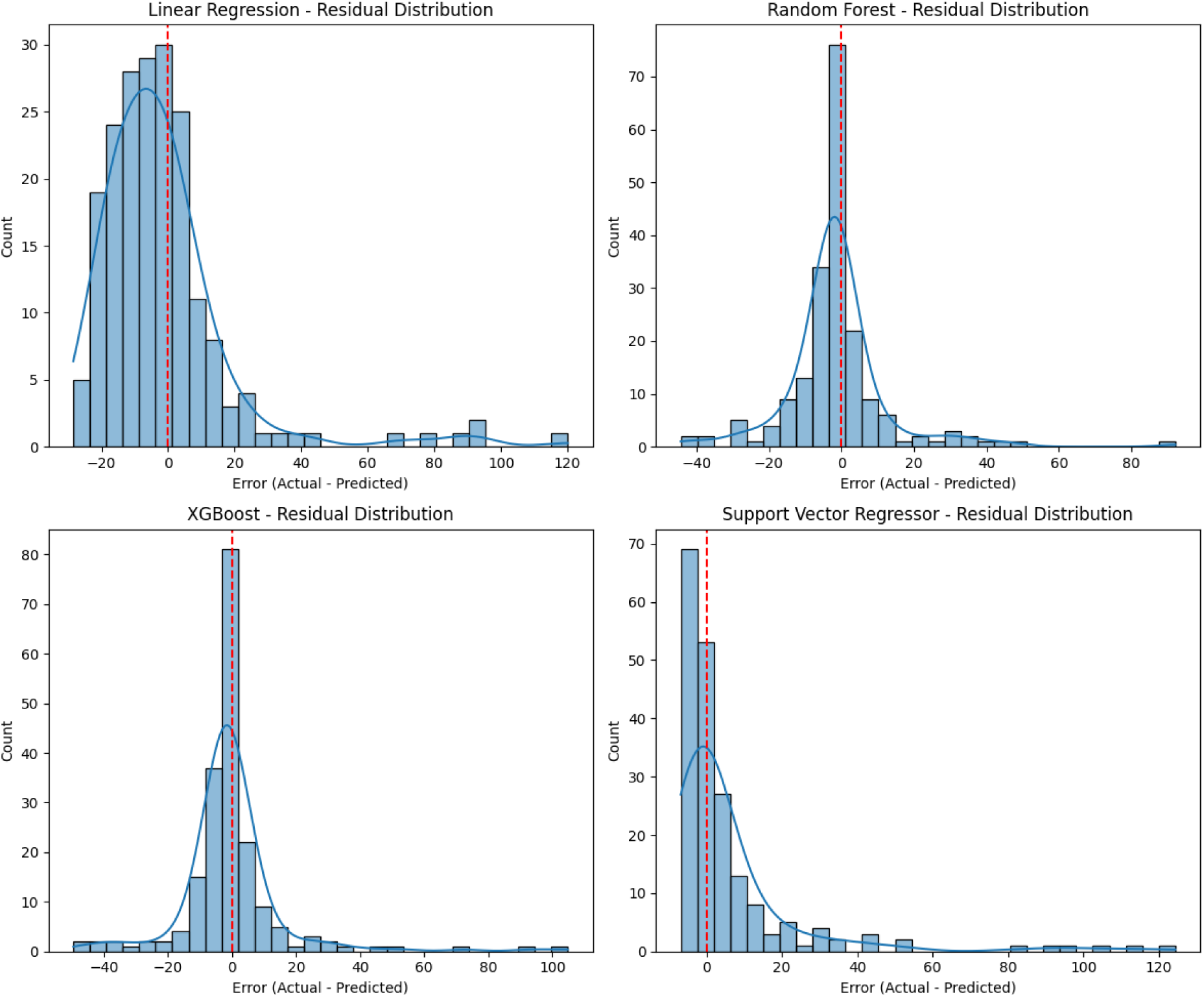

Correlation analysis among input variables showed low multicollinearity and weak linear associations, supporting the choice of a non-linear modelling approach.

**Figure 9.**
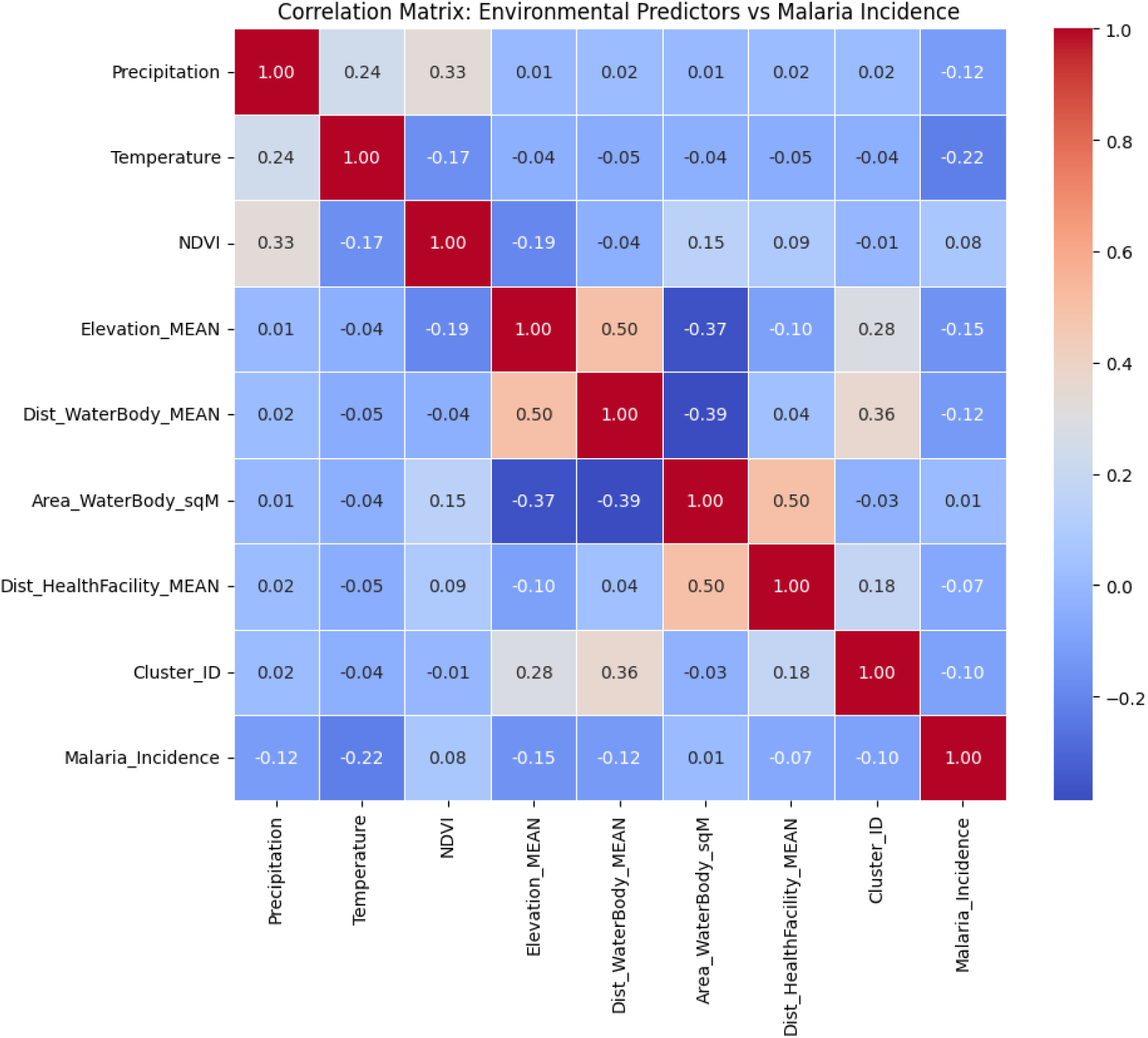

### Web Application Deployment and Interface

The *MalariaDash* prototype was successfully deployed as a Django-based web application and is available here – https://malariadash.kms.co.zw. It integrates satellite-derived data and machine learning outputs to generate malaria risk forecasts based on user-selected geographic coordinates and time frames.

**Figure 10.**
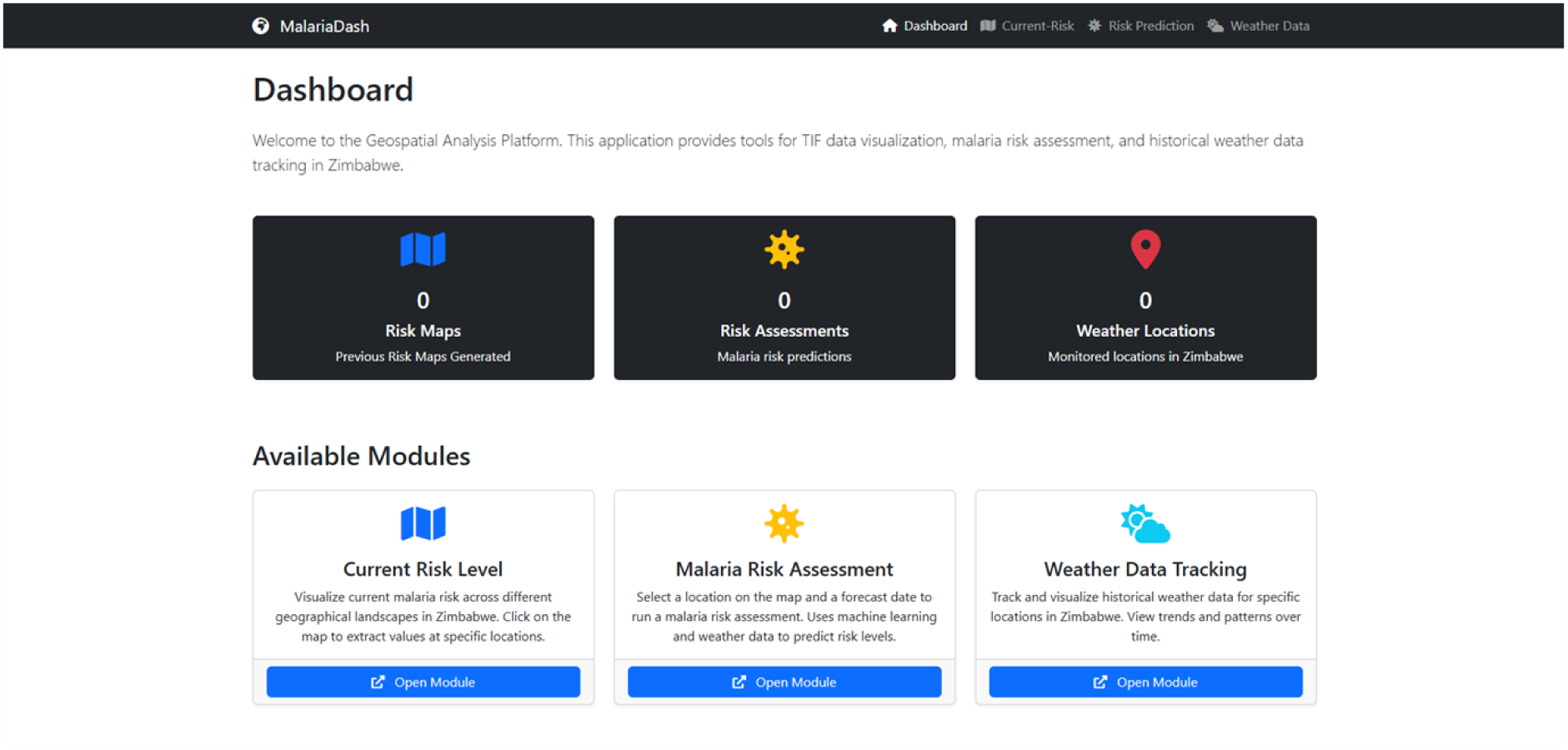

Users interact with the system via an intuitive map interface that allows the selection of prediction zones and dates. Upon submission, the system retrieves forecasted environmental data from the Weather API accessed through the Rapid API platform and computes malaria risk using the trained model. The output includes a predicted case count, a spatial marker and the predicted environmental metadata used in the prediction at the selected location.

**Figure 11.**
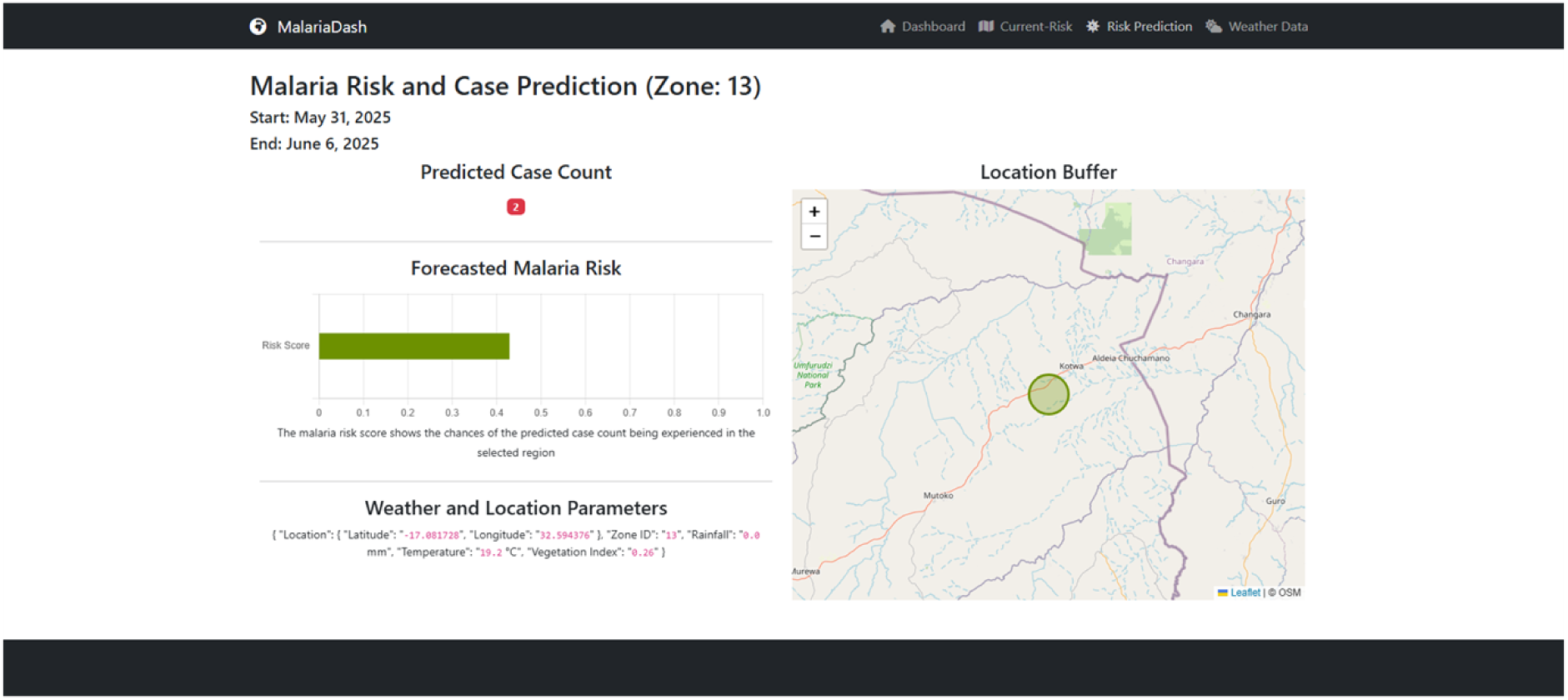

To support both periodic retraining and continuous improvement of the model over time, each prediction generated by the system is logged together with the associated input variables

### Seasonal Malaria Risk Mapping

Seasonal malaria risk maps produced via weighted overlay modelling in ArcGIS Pro revealed distinct spatial and temporal risk patterns as follows:

Table??

|  |  |
| --- | --- |
| Wet Season | Broadly distributed risk, with hotspots in Clusters 25, 27, 30, and 22. Risk aligned with high precipitation and water body proximity. |
| Hot-Dry Season | Concentrated high risk in Clusters 12, 24, 25, and 30, likely reflecting residual water sources and favourable thermal conditions. |
| Cool-Dry Season | General suppression of risk, with persistent moderate risk in Ward 18 and adjacent zones. |

**Figure 12.**
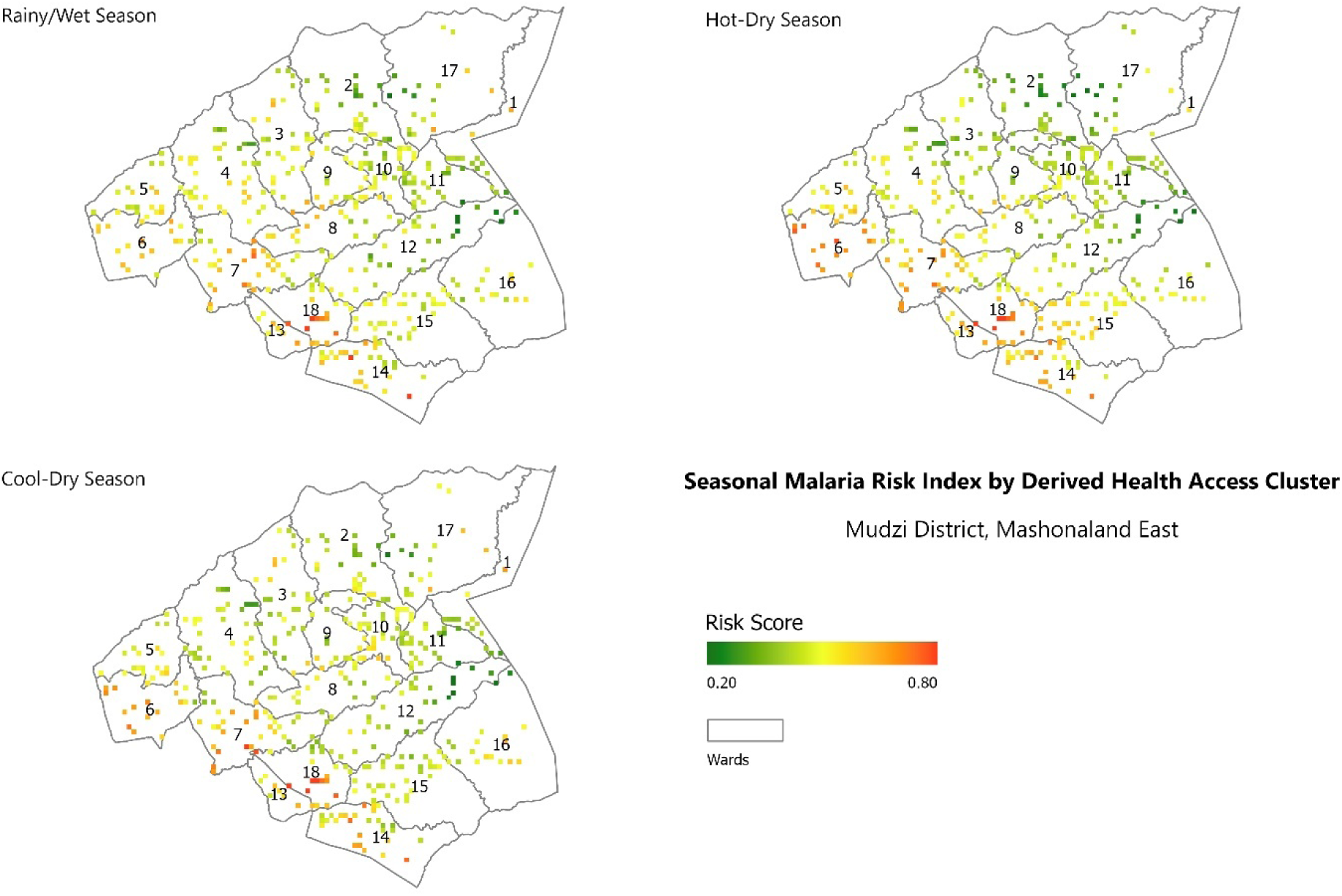
Malaria risk by derived health access cluster.

Cluster-level mapping provided finer spatial resolution, while ward-based stratification aligned with public health administrative units. Ward 18 consistently appeared as a high-risk zone across all three seasons.

**Figure 13.**
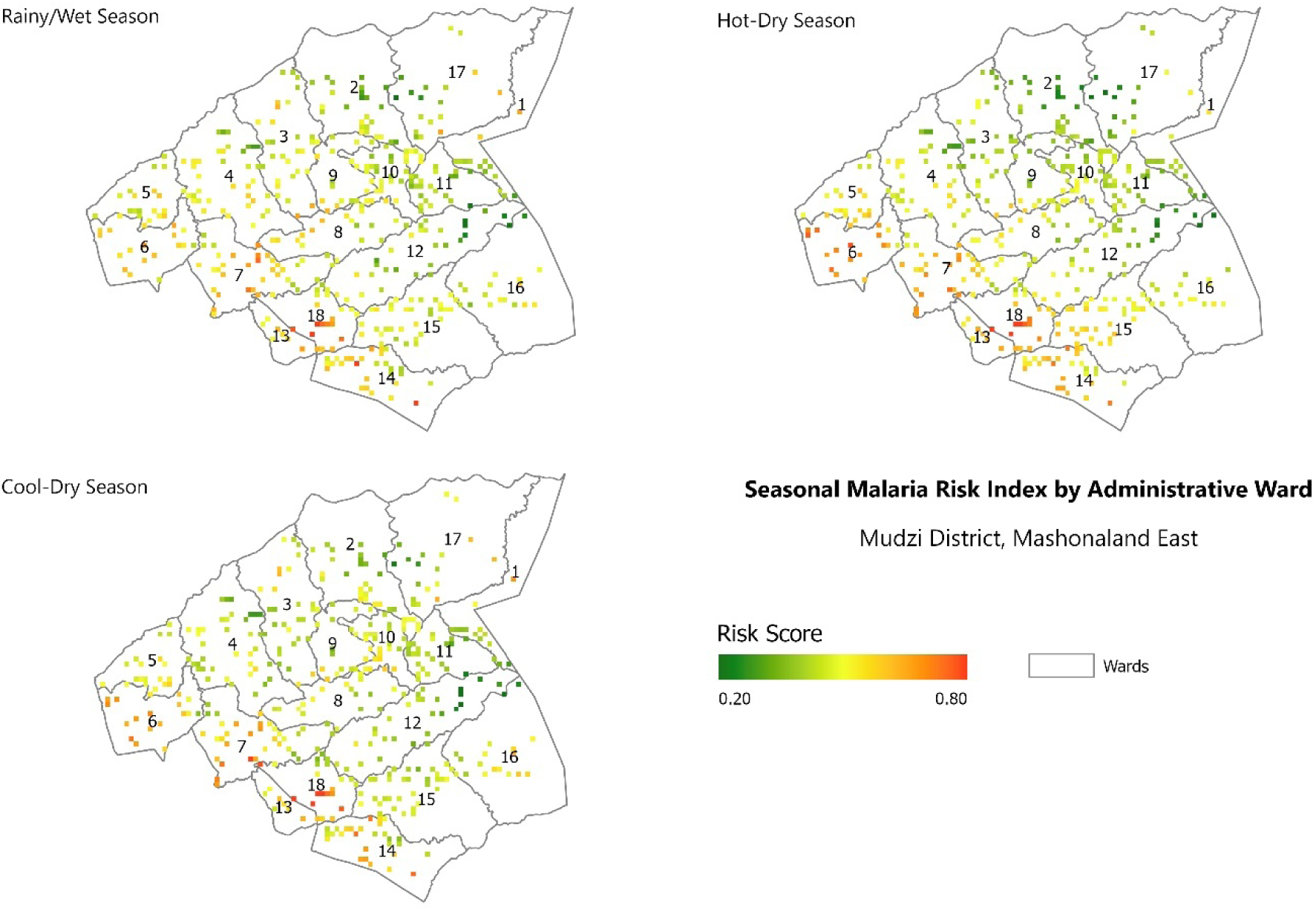
Malaria risk by administrative zone.

## Discussion and Conclusions

### Environmental Predictors and Temporal Effects

This study confirms that environmental variables play a critical role in shaping malaria transmission dynamics in rural Zimbabwe. Land surface temperature, proximity to stagnant water, and access to healthcare emerged as consistent predictors of malaria incidence. Notably, temperature demonstrated a statistically significant relationship with case counts both directly and at a two-month lag, aligning with established entomological knowledge of the Anopheles mosquito life cycle and Plasmodium development period (12). The inclusion of lagged environmental variables strengthened model performance and emphasized the temporally delayed influence of ecological factors on transmission.

The mixed results for NDVI further highlight the complexity of vector-environment relationships. While vegetation greenness did not achieve significance in isolation, it gained interpretive strength through its interaction with elevation. This points to the importance of accounting for terrain-mediated microclimatic variation when assessing habitat suitability for vector species.

### Machine Learning for Localised Prediction

The Random Forest model used in this study proved effective in capturing the non-linear relationships among environmental variables and malaria incidence. Unlike linear models, it accounted for variable interactions and produced stable predictions across the full range of incidence values. With an R² of 0.502 and the lowest RMSE among the tested models, it met the threshold for operational use in a rural district setting where hyper-local decision-making is essential.

Residual plots and correlation analyses supported the robustness of the model, with minimal bias and no problematic multicollinearity. This reinforces the utility of ensemble methods in low-resource contexts where data quality may vary, and linear assumptions are rarely met.

### MalariaDash Web-Application

The deployment of the *MalariaDash* application represents a practical transition from statistical modelling to decision-support infrastructure. By linking predictive algorithms to real-time satellite data through an intuitive web interface, the tool enables local health officers to forecast malaria risk by location and time window. This capability addresses a persistent weakness in Zimbabwe’s malaria surveillance system namely, its reliance on reactive paper-based reporting that lags behind epidemic spread.

The modular structure of the system supports future extensibility, such as integration with national health information platforms or the addition of mobile interfaces for field deployment.

### Seasonal Mapping for Strategic Intervention

The seasonal malaria risk maps provided an additional spatial-temporal layer for decision-making. By stratifying risk across both administrative wards and health access clusters, the maps offered a dual-scale view of vulnerability that is useful for both resource targeting and epidemiological understanding. Ward 18 consistently emerged as a high-risk zone, suggesting the presence of persistent structural and ecological risk factors.

Notably, the hot-dry season produced the most concentrated high-risk zones, despite lower rainfall. This reinforces findings from prior research that residual breeding habitats and sustained temperatures can prolong transmission even outside traditional peak seasons. The weighted overlay method used in ArcGIS, guided by statistical model outputs, ensured that risk classification was grounded in empirical significance rather than arbitrary thresholds.

This study demonstrates the feasibility and value of integrating environmental intelligence with machine learning and web technologies for malaria forecasting in low-resource settings. By operationalising a predictive model into a real-time web application *MalariaDash* the research contributes a functional tool for supporting precision vector control at the sub-district level in Zimbabwe.

### Recommendations

Based on the findings from Mudzi District, it is recommended that predictive systems like *MalariaDash* be integrated with national surveillance platforms such as DHIS2 to enable real-time data exchange and continuous model refinement. The framework should be adapted for use in other high-incidence districts, with recalibration using locally validated environmental data. To ensure adoption, capacity building for environmental health officers is essential, focusing on spatial data interpretation and digital tool usage. Periodic retraining of the model is also advised to maintain accuracy as environmental patterns evolve. Lastly, future research should incorporate sociodemographic variables such as income, housing conditions, and preventive practices to enhance the model’s specificity and broaden its relevance for intervention planning.

## Data Availability

The datasets used and/or analysed during the current study are not publicly available owing to restrictions by the Ministry of Health and Child Care but may be available from the corresponding author upon reasonable request.

## Acknowledgments and Funding

The authors gratefully acknowledge the Ministry of Health and Child Care (MoHCC) of Zimbabwe for providing access to health facility data and for authorising the conduct of the research in Mudzi District. The European Space Agency (ESA), through the Network of Resources (NoR) initiative, is thanked for providing technical sponsorship and access to cloud-based satellite imagery resources.

No additional external funding was received for the development or deployment of the *MalariaDash* web application.

## Appendix

**Figure 14.**
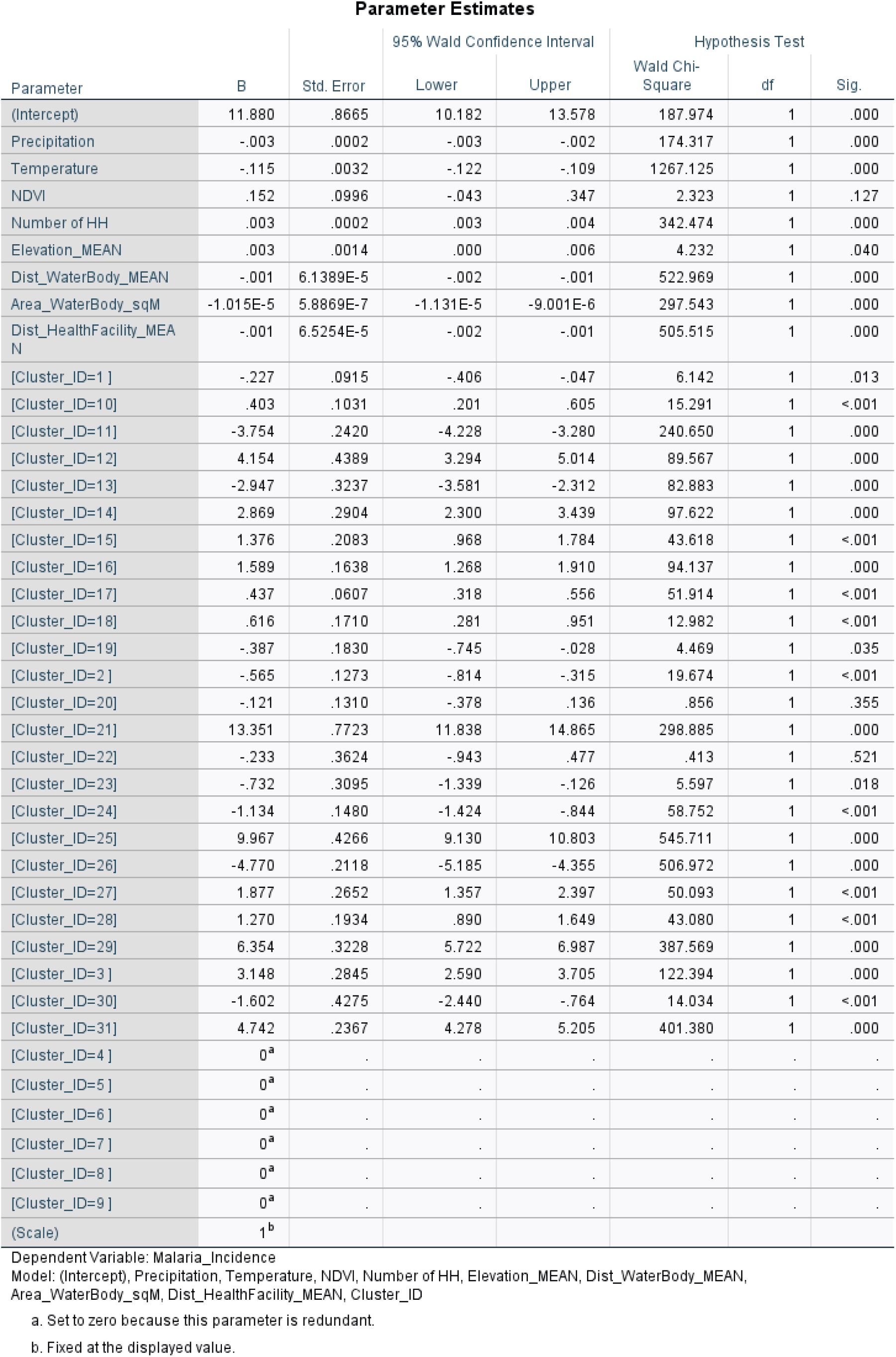
Parameter estimates for the Poisson model.

**Figure 15.**
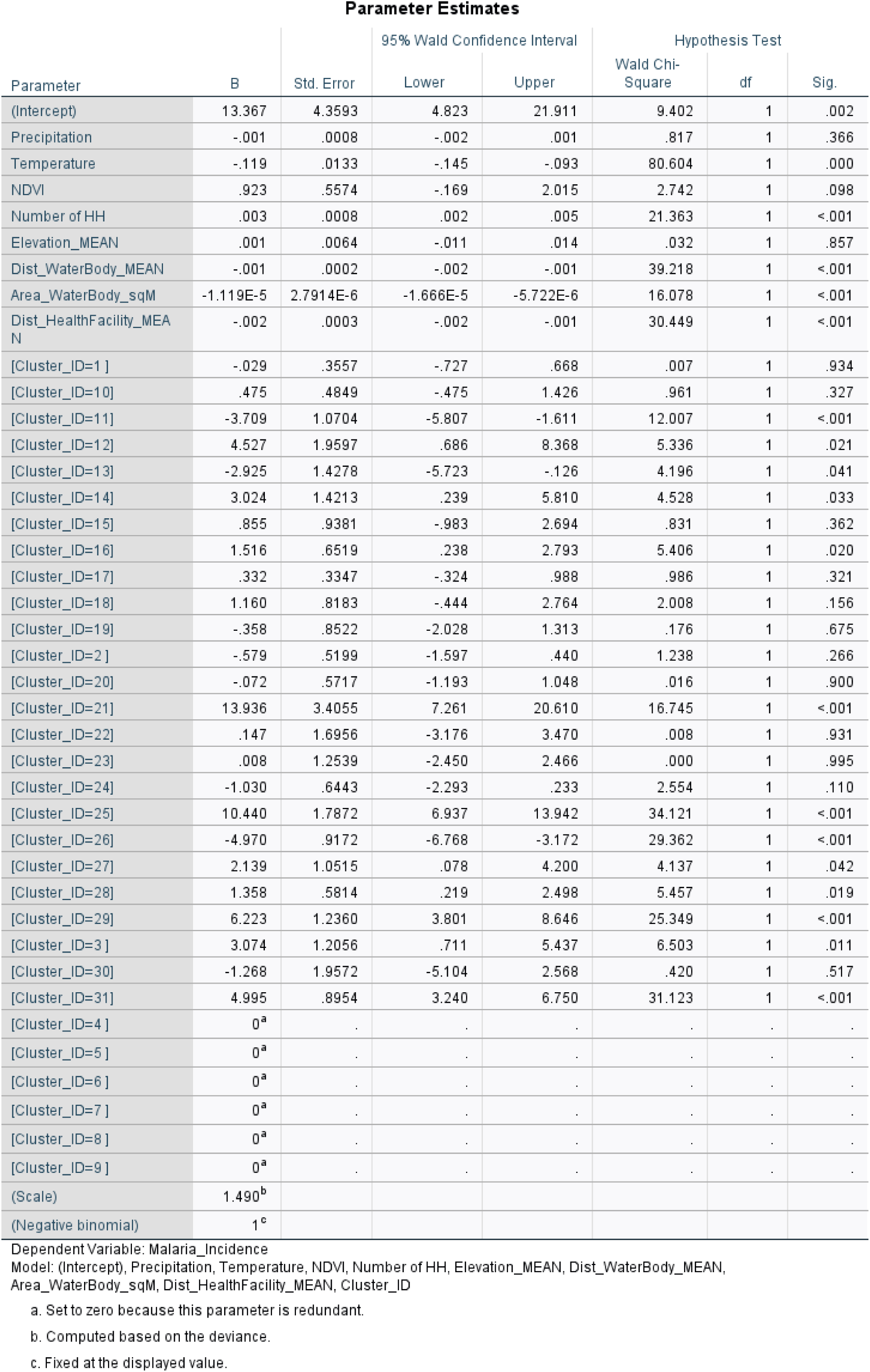
Parameter estimates for the Negative Binomial Model.

## References

1. Regional Data and Trends Briefing Kit. World Health Organisation; 2024 Dec. (World Malaria Report).

2. Corbel V, N’Guessan R, Corbel V, N’Guessan R. Distribution, Mechanisms, Impact and Management of Insecticide Resistance in Malaria Vectors: A Pragmatic Review. In: Anopheles mosquitoes - New insights into malaria vectors [Internet]. IntechOpen; 2013 [cited 2025 Jun 30]. Available from: https://www.intechopen.com/chapters/43899

3. Ministry of Health and Child Care of Zimbabwe. The National Health Profile 2020 Report. Harare: Government of Zimbabwe; 2020.

4. Ministry of Health and Child Care. Mashonaland East 2023 Provincial Health Report. Mashonaland East; 2023.

5. Food and Nutrition Council, World Food Programme. Mudzi District Food and Nutrition Security Profile. Mudzi District Office: Government of Zimbabwe; 2022.

6. Mutapure S, Dhliwayo P, Juru T, Mandozana G, Shambira G, Gombe N, et al. Malaria incidence in Zimbabwe, 2021: A secondary data analysis. J Interv Epidemiol Public Health [Internet]. 2025 Mar 20 [cited 2025 Jun 30];8(9). Available from: https://www.afenet-journal.net/content/article/8/9/full/

7. Bationo CS, Gaudart J, Dieng S, Cissoko M, Taconet P, Ouedraogo B, et al. Spatio-temporal analysis and prediction of malaria cases using remote sensing meteorological data in Diébougou health district, Burkina Faso, 2016–2017. Sci Rep. 2021 Oct 8;11(1):20027.

8. Gebreslasie MT. A review of spatial technologies with applications for malaria transmission modelling and control in Africa. Geospatial Health [Internet]. 2015 Nov 26 [cited 2025 Mar 15];10(2). Available from: https://www.geospatialhealth.net/gh/article/view/328

9. Zimbabwe National Statistics Agency. Zimbabwe Population and Housing Census Vol. 1. Zimbabwe: ZIMSTATS; 2022.

10. Funk C, Peterson P, Landsfeld M, Pedreros D, Verdin J, Shukla S, et al. The climate hazards infrared precipitation with stations—a new environmental record for monitoring extremes. Sci Data. 2015 Dec 8;2(1):150066.

11. Hierink F, Boo G, Macharia PM, Ouma PO, Timoner P, Levy M, et al. Differences between gridded population data impact measures of geographic access to healthcare in sub-Saharan Africa. Commun Med. 2022 Sep 16;2(1):1–13.

12. Agusto FB, Gumel AB, Parham PE. Qualitative assessment of the role of temperature variations on malaria transmission dynamics. J Biol Syst. 2015 Dec;23(04):1550030.

